# Increases in Organ Donation in Donor Hospitals Changing Organ Procurement Organization

**DOI:** 10.64898/2026.03.11.26348191

**Authors:** Isabella Sharifi, Erin Tewksbury, Matthew Wadsworth, David S Goldberg

## Abstract

**Importance:** Donor hospitals in the United States are assigned to a designated organ procurement organization (OPO) responsible for managing deceased donors in the designated donation service area (DSA). Donor hospitals can apply for waivers to work with a different OPO with appropriate justification, and beginning with the 2026 OPO certification cycle, the highest-performing OPOs can bid to work with donor hospitals managed by intermediate- and low-performing OPOs.

**Objective:** We sought to evaluate the impact of donor hospital waivers on organ donation activity.

**Design:** Retrospective cohort study.

**Setting:** We evaluated Organ Procurement and Transplantation Network (OPTN) data from two OPOs (Donor Network West and Honor Bridge), each with a donor hospital (Renown Regional Medical Center and North Carolina Baptist Hospital) in its DSA granted a waiver to work with a different OPO beginning in April 2025.

**Main Outcome:** We assessed changes in the number of organ donors and organs transplanted pre- and post-granting of a waiver using a difference-in-differences approach based on multilevel mixed-effects models.

**Results:** After switching OPO affiliations, these two donor hospitals had marked and statistically significant increases in the number of donors recovered and organs transplanted, despite stable numbers of reported deaths at each hospital. In multivariable models, switching OPO affiliations was associated with a statistically significant increase in donors recovered and organs transplanted. Conclusion: With eight months of post-waiver data, donor hospitals with granted waivers had significant increases in donation activity driven by improved donor conversion rather than changes in referral patterns or organ yield per donor. Although longer-term data are needed to confirm these findings, CMS and the organ transplant community should feel confident that changing donor hospital-OPO affiliations will not negatively impact donation and may lead to significant increases in donation. These data also counter unfounded concerns that the continued granting of waivers and realignments of donor hospital-OPO affiliations during the 2026 recertification cycle will lead to a collapse of the system of organ donation.

**KEY POINTS:** *Question:* Do donor hospitals who request a waiver to change OPO affiliations have changes in organ donation rates?

*Findings:* Using a difference-in-difference approach, the two donor hospitals who changed OPO affiliations had a significant increase in organ donors and organs transplanted after being granted a waiver.

*Meaning:* Donor hospitals that change OPO affiliations have an immediate increase in organ donation activity.

## INTRODUCTION

The approval of a new Centers for Medicare and Medicaid Services (CMS) organ procurement organization (OPO) final rule in 2020 led to increased oversight and monitoring of OPO performance by policymakers and Congress.^1^ The 2020 CMS OPO final rule allows the highest-performing OPOs (Tier 1) to compete for donor hospitals in the donation service areas (DSAs) of intermediate and low-performing OPOs (Tier 2 and Tier 3).^2^ However, there is also a pathway by which a donor hospital can request a CMS waiver to work with an OPO other than its designated OPO provided if the waiver is expected to increase organ donation.^2^

Despite being available since before 2006, the waiver pathway has rarely been requested until recently.^3-5^Concerns have been raised that waivers and changes in donor hospital affiliations could negatively impact organ donation in a given geographic area by ending longstanding donor hospital-OPO relationships. However, there are no empiric data to support this claim.

In April 2025, two large hospitals, Renown Regional Medical Center in Nevada and North Carolina Baptist Hospital changed OPO affiliations after being granted CMS waivers. Renown Regional Medical Center realigned from Donor Network West (CADN) to Nevada Donor Network (NVLV) and North Carolina Baptist Hospital from Honor Bridge (NCNC) to LifeShare Carolinas (NCCM). Both realignments were subsequently met with legal challenges, and recently, several other waiver applications, all seeking to switch affiliations to NCCM.^6-11^

Concerns about the granting of waivers, along with the prospect of increased competition and donor hospitals switching affiliations after the 2026 OPO certification cycle has magnified the need to empirically study the impact of donor hospitals realigning with different OPOs.

## METHODS

We evaluated Organ Procurement and Transplantation Network (OPTN) data from August 1, 2024 through November 30, 2025 to ensure equal time pre-versus post-OPO waivers. Within each DSA, the OPTN provided monthly aggregate data per donor hospital on deaths reported to the OPO, donors recovered, and organs transplanted. We focused on two OPOs (CADN and NCNC) to compare organ donation activity at the hospitals that maintained their affiliation (i.e., “control”) to the two hospitals that changed OPO affiliation (i.e., “intervention”).

### Statistical analyses

We first evaluated the monthly number of donors recovered and organs transplanted for control versus intervention donor hospitals. We employed a difference-in-differences (DID) analytic framework to evaluate the association between hospitals changing OPO affiliation and hospital-level donor activity.^12^ This approach allowed us to estimate the causal impact of an intervention (hospitals switching OPO affiliations) by comparing the change in outcomes (monthly donors recovered and organs transplanted) between treatment group (hospitals switching affiliations) and control group (hospitals maintaining their affiliation).

The primary outcomes were: a) donors recovered per month; and b) organs transplanted per month. We fit multilevel mixed-effects models (using continuous outcomes) with random intercepts for each hospital. This method accounted for differences in baseline number of donors recovered and organs transplanted for each hospital and to account for within-hospital correlations. Models included the covariate of calendar month, a variable for era pre-post OPO waiver implementation (8/1/2024-3/31/2025 versus 4/1/2025-11/30/2025), and an interaction term between waiver implementation period and control versus intervention hospital. This interaction term represented the DID estimator and signified whether donor hospitals changing OPO affiliations were associated with a difference in organ donation activity. A p-value of <0.1 was considered statistically significant for the interaction term.^13^

This study was considered exempt from Institutional Review Board review.

## RESULTS

### Donors recovered

The mean number of monthly donors recovered at control hospitals was unchanged in the post-waiver period: 56.6 versus 57.5, although there was some month-to-month variation (Figure 1a). In contrast, mean monthly donors increased at the intervention hospitals after switching OPO affiliations (mean donors per month: 5.6 [pre-waiver] versus 7.9 [post-waiver]). These increases were evident at both intervention hospitals.

**Figure 1.**
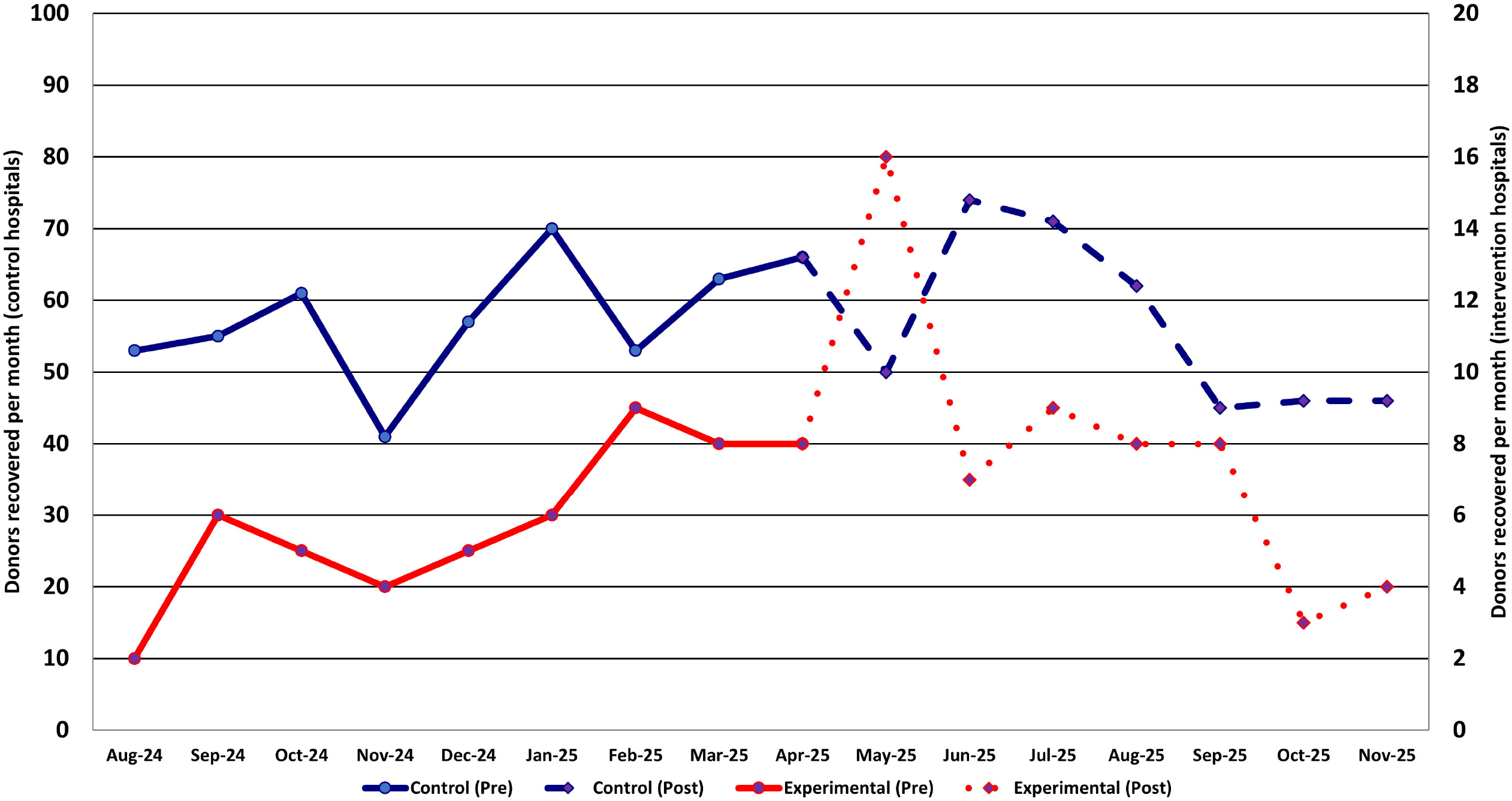
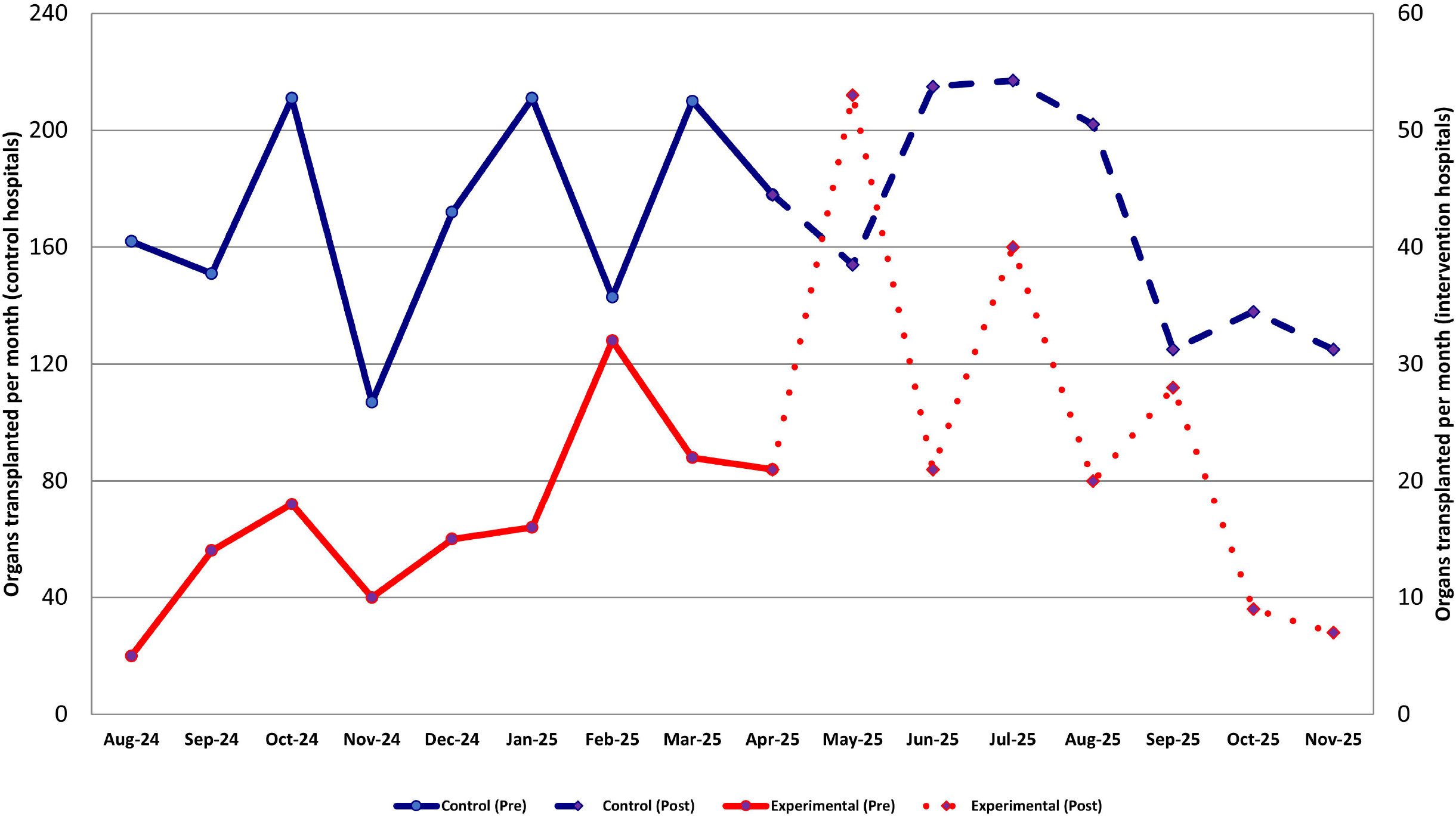
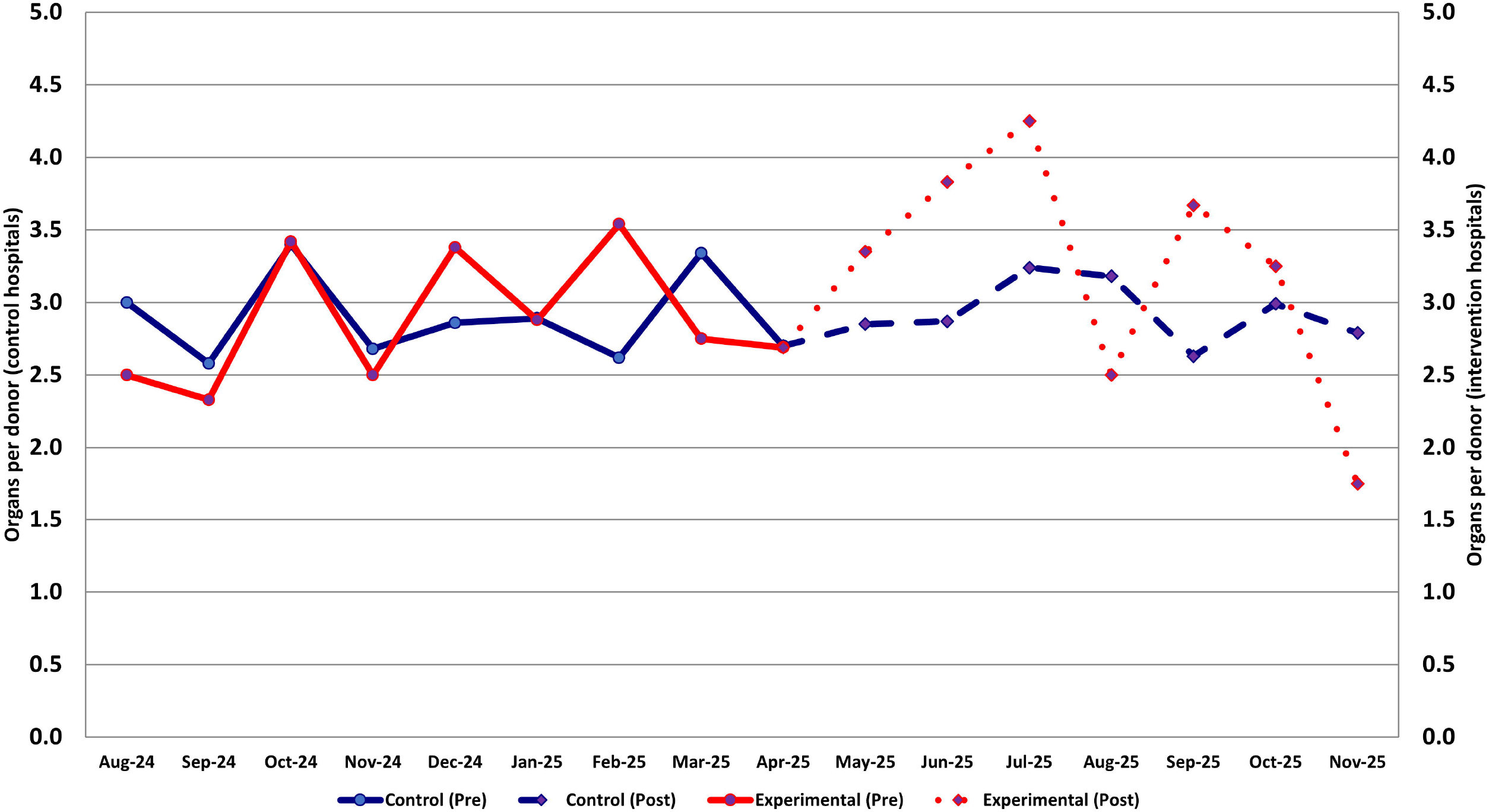
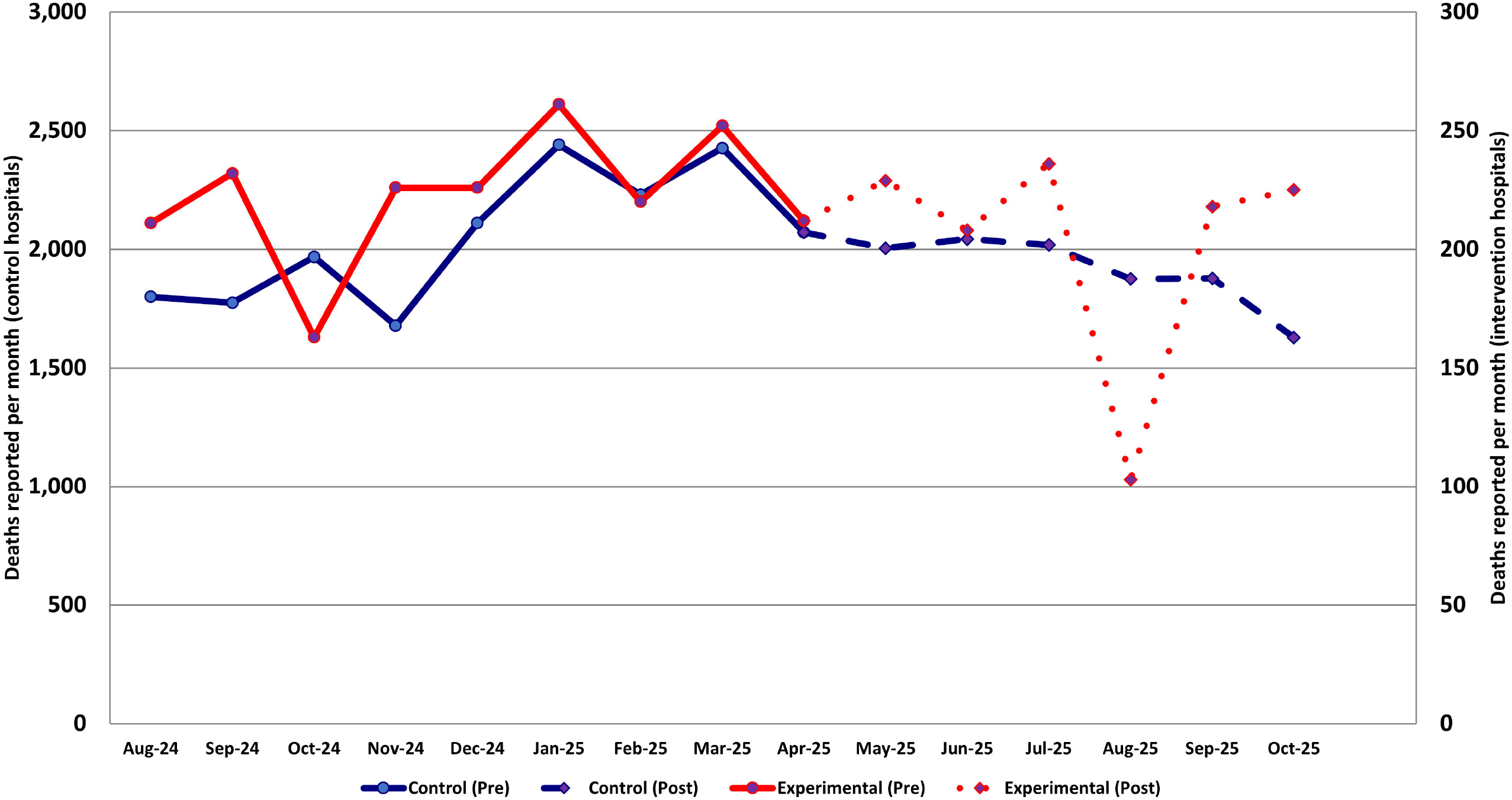
(four panels): Monthly organ donation activity at control and intervention hospitals in the two analyzed donation service areas from 8/1/2024-11/30/2025. **a**. Figure 1a: Donors recovered **b**. Figure 1b: Organs transplanted **c**. Figure 1c: Organs transplanted per donor **d**. Figure 1d: Deaths reported by donor hospitals to the Organ Procurement and Transplantation Network **e**. *Note: The intervention hospitals are Renown Regional Medical Center and North Carolina Baptist Hospital which were each granted a waiver to work with a different OPO. The control hospitals are all other donor hospitals within the DSA of Donor Network West and Honor Bridge. Dashed lines (control) and dotted lines (intervention) with diamond data points reflect the post-waiver period of 4/1/2025–11/30/2025 for all hospitals. Deaths reported not available for November 2025 for all hospitals so it was excluded.

### Organs transplanted

The mean number of organs transplanted per month at control hospitals was unchanged in the post-waiver period (170.9 pre-waiver versus 169.2 post-waiver) but increased substantially at the intervention hospitals (16.5 pre-waiver versus 24.9 post-waiver; Figure 1b). Changes were similar at both intervention hospitals. Overall mean number of organs transplanted per donor was stable at intervention hospitals, and increased at intervention hospitals (Figure 1c), but was limited to Renown Regional Medical Center (3.2 pre-waiver versus 3.6 post-waiver).

### Reported deaths

Despite month-to-month fluctuation in the number of reported deaths, the mean number in control and intervention hospitals was similar in the pre-versus post-waiver periods (Figure 1d).

### Multivariable models

In mixed-effects linear regression models, the difference-in-difference estimator (interaction of control versus experimental donor hospital x waiver period) was significant (p<0.1) in both models—the number of donors recovered and organs transplanted per month changed significantly more in the experimental hospitals in the post-waiver period than in control hospitals.

### Changes in practice with donor hospital changing OPO affiliation

To explore potential mechanisms by which a donor hospital changing OPO affiliations could be associated with changes in donor activity, we communicated with leadership from LifeShare Carolinas. Several changes in practice were identified:

- LifeShare and North Carolina Baptist Hospital created leadership committees to address areas where donation processes were inefficient.
- LifeShare clinical, family services, and hospital development staff are onsite and available to hospital staff on a daily basis.
- LifeShare implemented electronic referrals and communication between OPO and donor hospital staff using Epic Secure Chat.
- LifeShare invested heavily in their Family Services division, with additional investments in preparation for adding a donor hospital.

## DISCUSSION

We demonstrate that in the short-term period after two large hospitals changed OPO affiliations, there were significant increases in the number of donors recovered and organs transplanted despite an unchanged number of reported deaths. These data suggest that donor hospital waivers are not harmful and may potentially lead to an increase in donor activity, and lives saved through organ transplantation.

Previous studies have demonstrated that data-driven performance improvement initiatives can lead to a rapid and significant increase in organ donation.^14^ Furthermore, there are data to demonstrate that while OPO leadership changes do not guarantee improvements in organ donation, they significantly increase the odds of rapid improvement.^15^ Our data validate these prior studies, but for the first time, examine a the potential for improvements in organ donation at individual hospitals simply by virtue of changing OPO affiliations. The data we present from LifeShare Carolinas help to explain how these rapid improvements can occur. OPOs have heterogeneous practices, and higher-performing OPOs may implement changes that are specific to the new donor hospital, or broadly implemented across the OPO, that provide an opportunity for marked and immediate improvements in organ donation.

We recognize that we only evaluated data from two donor hospitals at two OPOs, with eight months of follow-up after their acquisition. Longer-term data are needed to evaluate the durability of these improvements, and whether they always occur when there is a change in affiliation of a donor hospital and an OPO. Nevertheless, these data are time-sensitive and vital to the transplant and donation community, and government regulators in light of the 2026 OPO certification cycle and the potential for future donor hospital waiver requests to CMS. In addition, these data underscore that donor hospital acquisitions may have the potential to improve organ donation and should lead CMS to consider future waivers based on these encouraging preliminary data.

## Data Availability

The raw and aggregated data cannot be shared as per the data use agreements of the OPTN, but if an investigator obtains the data we can share our underlying source code.

## Abbreviations

(CMS): Centers for Medicare and Medicaid Services
(OPO): Organ procurement organization
(DSA): Donation service areas
(CADN): Donor Network West
(NVLV): Nevada Donor Network
(NCNC): Honor Bridge
(NCCM): LifeShare Carolinas
(DID): Difference-in-differences

## Acknowledgments

This work was supported in part by Health Resources and Services Administration contract HHSH250-2019-00001C. The content is the responsibility of the authors alone and does not necessarily reflect the views or policies of the Department of Health and Human Services, nor does mention of trade names, commercial products, or organizations imply endorsement by the U.S. Government. We would like to thank Michael Daniels, Executive Director of LifeShare Carolinas and Dr. Michael Haley, Medical Director of LifeShare Carolinas for providing process-level information related to changes in donor management after North Carolina Baptist Hospital began to work with their OPO. Michael Daniels and Dr. Michael Haley reviewed the manuscript to ensure the information related to changes in practices at North Carolina Baptist Hospital were accurate, but they did not provide input on the OPTN data analyses or the conclusions of the manuscripts.

